# The Safety and Efficacy of Transcatheter Edge-to-Edge Mitral Valve Repair for Central versus Non-central Primary Mitral Regurgitation: A Single-center Cohort Study

**DOI:** 10.1101/2025.06.02.25328830

**Authors:** Yusuke Enta, Masaki Miyasaka, Yoshiko Munehisa, Daishi Tazawa, Yuhei Hasegawa, Momo Kosuga, Manabu Maeda, Masatoshi Sato, Yun Teng, Natsuko Satomi, Makoto Saigan, Yuta Kobayashi, Masaki Nakashima, Yusuke Toki, Yukihiro Hayatsu, Masaki Hata, Ayumi Nojiri, Sae Ochi, Norio Tada

**Affiliations:** Department of Cardiovascular medicine, Sendai Kousei Hospital, 1-20 Tsutsumidori-amamiyamachi, Aoba-ku, Sendai, Miyagi 9810914, Japan; Department of Laboratory Medicine, The Jikei University School of Medicine, 3-19-18 Nishi-Shimbashi, Minato-ku, Tokyo 1058471, Japan; Department of Cardiovascular surgery, Sendai Kousei Hospital, 1-20 Tsutsumidori-amamiya, Aoba-ku, Sendai, Miyagi 9810914, Japan

**Keywords:** Primary mitral regurgitation

## Abstract

**Background:** It remains unclear whether transcatheter edge-to-edge mitral valve repair (M-TEER) ensures procedural safety and 30-day and one-year efficacy in non-central primary mitral regurgitation (pMR) compared to central pMR.

**Objectives:** This study assessed clinical and echocardiographic outcomes of M-TEER in non-central pMR compared with central pMR.

**Methods:** This study included consecutive patients with pMR who underwent M-TEER at a single center between April 2018 and December 2022. Data were prospectively collected and analyzed retrospectively. Baseline clinical and echocardiographic characteristics, as well as procedural and clinical outcomes up to 1-year post procedure, were compared between patients with non-central pMR group (NCpMR) and those with central pMR group (CpMR).

**Results:** Among 129 patients with pMR who underwent M-TEER, 50 had NCpMR and 79 had CpMR. Neither group had 30-day mortality. Although residual MR grade 3+/4+ was more frequent in NCpMR than CpMR (10.0% vs. 0.8%, p = 0.01), MR worsening was lower in NCpMR at 6 months (5.0% vs. 10.5%, p = 0.18) and 12 months (7.5% vs. 27.7%, p = 0.03). A 30-day landmark analysis showed a trend towards a lower prevalence of residual MR grade 3+/4+ at 1 year in NCpMR (p = 0.051).

**Conclusions:** No significant differences in 30-day mortality and emergent intervention for complications were noted among two groups. Although MR grade at discharge was better in CpMR, NCpMR showed lower MR worsening and sustained MR reduction at 1-year. M-TEER can be an option for non-central pMR in high-risk surgical patients.

## Introduction

Severe mitral regurgitation (MR) is associated with a significant increase in morbidity and mortality^1–4^. Mitral valve (MV) surgery remains the standard treatment for symptomatic patients with primary MR (pMR)^5^. However, nearly half of the patients referred for surgery^6,7^ do not undergo the procedure, primarily due to comorbidities such as left ventricular (LV) dysfunction or advanced age^8^. For these patients, Mitral transcatheter edge-to-edge repair (M-TEER), a catheter-based intervention, has emerged as an alternative treatment strategy. The M-TEER procedure, most commonly performed using the MitraClip system (Abbott Vascular, California, USA), replicates the surgical edge-to-edge MV repair technique^9–11^.

The Endovascular Valve Edge-to-Edge Repair Study (EVEREST) II trial, a pivotal study conducted in the United States (US), compared percutaneous MV repair using the MitraClip system with standard MV surgery. The trial demonstrated that although the MitraClip approach is less effective in terms of long-term durability, it offers a superior safety profile, with fewer complications and a faster recovery time. These findings established the safety and efficacy of M-TEER with the MitraClip for pMR. Based on these results, the US Food and Drug Administration (FDA) approved M-TEER for the treatment of pMR, making EVEREST II the first randomized controlled trial to support such regulatory approval.

In the early days of M-TEER, clinical trials included only patients with central pMR, defined as MR originating from structural alterations involving the central area (A2/P2)^12^. This selection was based on the premise that a central regurgitant area is an important anatomic criterion for achieving optimal procedural outcomes^13^. When a non-central area is targeted, concerns arise regarding potential device interference with the subvalvular apparatus and challenges in image guidance, which could compromise the efficacy and safety of the procedure^14,15^. For instance, device entanglement with the subvalvular apparatus may lead to chordal rupture, increasing the risk of procedural failure and worsening MV function.

In real-world practice, following the commercial approval of M-TEER in the US, approximately 20% of patients undergoing the procedure have non-central pMR^12^. Therefore, it is essential to establish robust clinical evidence regarding the safety and efficacy of M-TEER for non-central area. However, only one original study, published in 2013, has specifically evaluated M-TEER in non-central pMR, including 79 patients and demonstrating similar short-term (up to 6 months) clinical outcomes between central and non-central pMR^16^. Consequently, evidence supporting the long-term efficacy and safety of M-TEER in non-central pMR remains limited. To address this gap, the present study compares the procedural and 1-year safety and efficacy of M-TEER with the MitraClip system between patients with central and non-central pMR.

## Methods

### Study Population and Data Collection

This retrospective analysis included patients with primary mitral regurgitation (pMR) who underwent M-TEER using the MitraClip system at Sendai Kousei Hospital (Sendai, Miyagi, Japan) between April 2018 and December 2022. Among 374 consecutive patients treated for symptomatic significant MR, only those diagnosed with pMR were included. All patients were deemed high surgical risk and selected for M-TEER by a multidisciplinary heart team. Clinical and procedural data were prospectively collected in an institutional M-TEER database, including patient demographics and echocardiographic findings (transthoracic echocardiography (TTE) and transesophageal echocardiography (TEE)) from the preoperative, intraoperative, postoperative, and follow-up periods.

### Assessment of MR

MR severity was graded using a multiparametric approach according to current guidelines with grades none/trace (0), mild (1+), mild-to-moderate (2+), moderate-to-severe (3+), and severe (4+) in TTE^17,18^. Significant MR was defined as 3+ or 4+ (3+/4+). The assessment was performed at baseline, discharge, and 30-day, 6-month, and 12-month follow-up. Preoperatively, all patients underwent evaluation using transesophageal echocardiography (TEE). MR severity was assessed by our facility’s echocardiologists using a comprehensive analysis of quantitative and semiquantitative echocardiographic parameters as recommend^19,20^. Central MR was defined as MR originating from the central portion of the coaptation line (A2/P2), whereas non-central MR was defined as MR originating from the lateral or medial segments of the coaptation line (A1/P1 and A3/P3), including the commissures (**Figure 1**).

**Figure 1.**
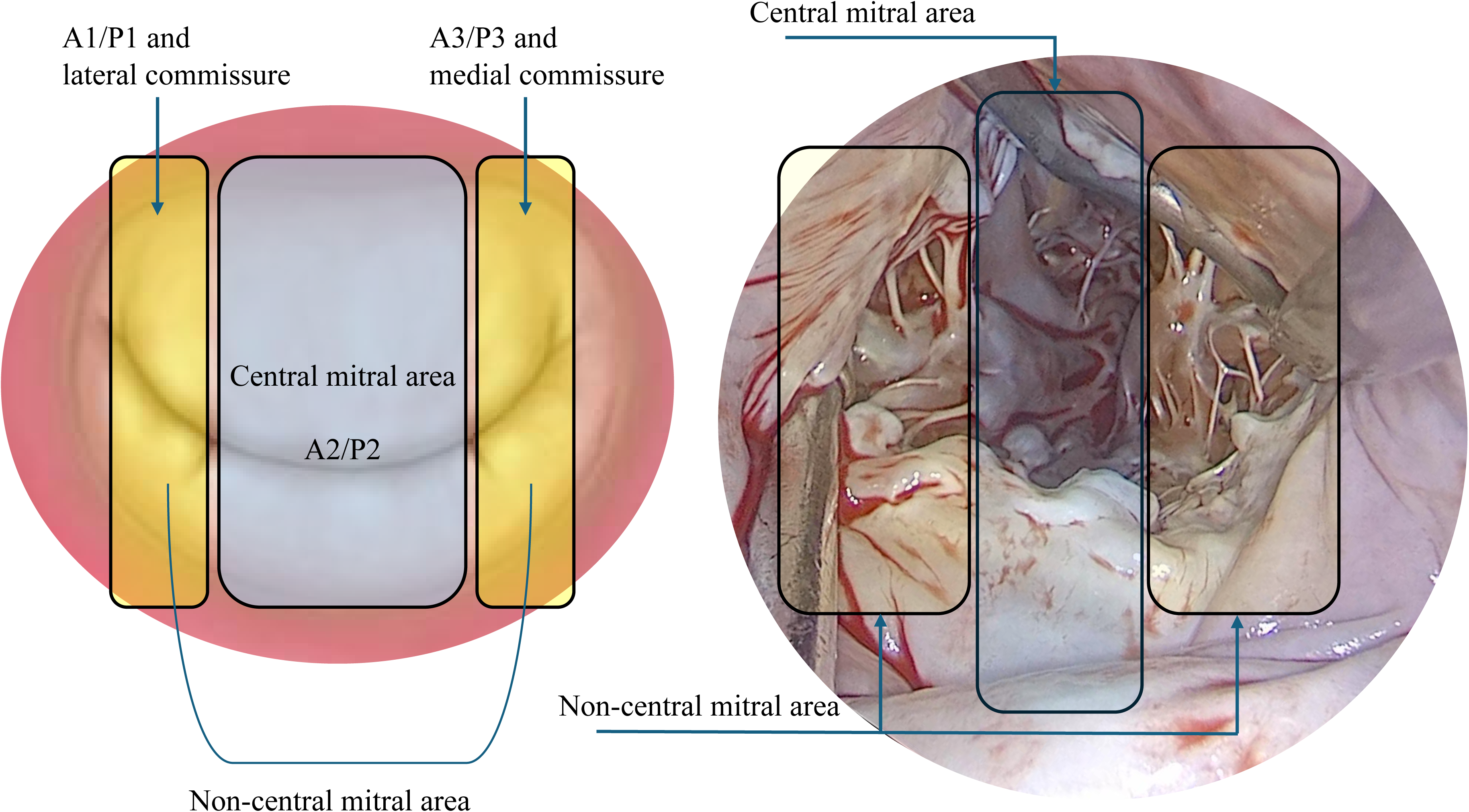
Definition and schematics of central and non-central mitral valves. The illustration depicts mitral valve anatomy, with central MV defined by the A2P2 scallop and non-central MV including A1P1, A3P3 scallops, and commissures.

### Study endpoints

The safety endpoints of the study included 30-day mortality and the rate of emergency surgery or reintervention related to the device or vascular access. The efficacy endpoints included residual MR grade at discharge, as well as at 30 days, 6 months, and 12 months post-procedure. Technical success and device success were retrospectively adjudicated by experienced cardiologists using the Mitral Valve Academic Research Consortium (M-VARC) criteria^21,22^. The variables analyzed in this study are listed in **Supplemental Table 1**. Composite events in the Kaplan-Meier curve were defined as deployment failure, single-leaflet device attachment (SLDA), and residual MR grade 3+/4+.

### Procedure and device

The M-TEER procedure using the MitraClip system has been described in detail elsewhere^9^. The MitraClip system transitioned from the second-generation (G2) to the fourth-generation (G4) system in September 2020.

### Criteria and clinical follow-up

Individual patient data were evaluated and monitored in accordance with the recommendations of the M-VARC^21^. Follow-up visits were scheduled at 30 days, 6 months, and 12 months after M-TEER to assess clinical outcomes and TTE findings.

### Statistical analysis

Continuous variables were assessed for normality using the Shapiro-Wilk test. Variables following a normal distribution were reported as the mean ± standard deviation (SD), while non-normally distributed variables were presented as the median and interquartile range (IQR). Student’s t-test was used for intergroup comparisons of parametric data, whereas the Mann-Whitney U test was applied for non-parametric data. Categorical variables were expressed as counts (percentages) and compared using Pearson’s χ² test or Fisher’s exact test, as appropriate. A p-value < 0.05 was considered statistically significant. The incidence of recurrent MR after M-TEER was estimated using the Kaplan-Meier method, and differences between groups were assessed with the log-rank test. All statistical analyses were performed using JMP V.14 for Mac (SAS Institute, North Carolina, USA).

### Ethical consideration

Our registry was approved by the local Ethical Committee at the Sendai Kousei Hospital in accordance with the Declaration of Helsinki (IRB Number 7–9). All patients provided written informed consent for participation in the registry.

## Results

### Baseline characteristics

A total of 129 patients with pMR were identified, with a mean age of 81.1 ± 8.7 years, and 61.2% were male. The central pMR group (CpMR) included 79 patients (61.2%), while the non-central pMR group (NCpMR) consisted of 50 patients (38.8%) (**Figure 2**). Among the NCpMR cases, A1/P1 lesions were identified in 17 patients, and A3/P3 lesions in 33 patients. Baseline characteristics are detailed in **Table 1**. The majority of patients presented with New York Heart Association (NYHA) functional class III or IV symptoms (64.0% vs. 50.6%, p = 0.45). Compared to CpMR, patients in the NCpMR had a significantly higher prevalence of atrial fibrillation (AF) (80.0% vs. 59.5%, p = 0.01) and a more frequent history of prior transcatheter structural heart disease interventions (10.0% vs. 0%, p<0.01). Additionally, the Society of Thoracic Surgeons (STS) score for MV replacement was higher in the NCpMR than in the CpMR (12.3 [7.3–15.0] % vs. 9.5 [5.9–11.8] %, p = 0.02). Pre- and post-procedural echocardiographic variables are summarized in **Table 2**. There were no significant differences in MR severity parameters, including regurgitant volume (RV) (99 ± 41 vs. 101 ± 43 mL, p = 0.83), regurgitant fraction (RF) (61 ± 18 vs. 64 ± 13%, p = 0.41), and effective regurgitant orifice area (EROA) (0.54 ± 0.25 vs. 0.60 ± 0.24 cm², p = 0.25).

**Figure 2.**
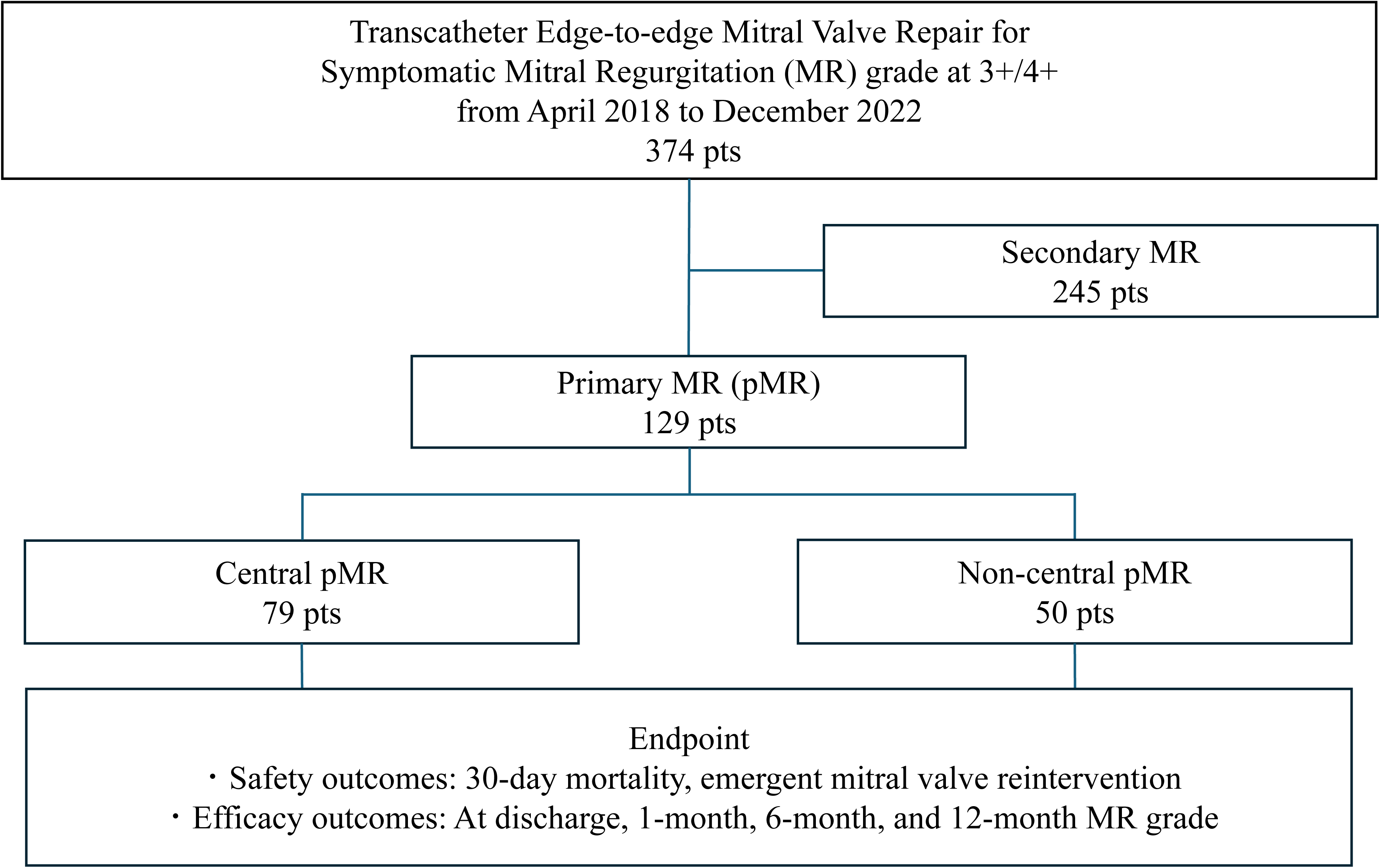
Study population, design, and endpoints. Among 374 consecutive patients treated between April 2018 and December 2022, 129 patients (34.5%) had primary mitral regurgitation (pMR). Of these, 79 (61.2%) were classified as central pMR, while the remaining 50 (38.8%) were classified as non-central pMR. Safety endpoints included 30-day mortality, emergency surgery, and reintervention. Efficacy endpoints assessed residual MR grade at discharge, 30 days, 6 months, and 12 months.

**Table 1.** Baseline characteristics.

|  | All<br>(n=129) | Non central<br>(n=50) | Central<br>(n=79) | p-value |
| --- | --- | --- | --- | --- |
| Age (y.o.) | 81.1±8.7 | 82.6±6.8 | 80.1±9.6 | 0.11 |
| Male, n(%) | 79(61.2) | 18(36.0) | 31(39.2) | 0.71 |
| HT, n(%) | 114(88.4) | 47(94.0) | 67(84.8) | 0.11 |
| DL, n(%) | 50(38.8) | 20(40.0) | 30(38.0) | 0.81 |
| DM, n(%) | 14(10.9) | 5(10.0) | 9(11.4) | 0.80 |
| AF/AFL, n(%) | 87(67.4) | 40(80.0) | 47(59.5) | 0.01 |
| Coronary artery disease, n(%) | 16(12.4) | 8(16.0) | 8(10.1) | 0.32 |
| Previous coronary intervention, n(%) | 15(11.6) | 7(14.0) | 8(10.1) | 0.51 |
| Prior open heart surgery, n(%) | 10(7.8) | 6(12.0) | 4(5.1) | 0.16 |
| Prior transcatheter SHD intervention, n(%) | 5(5.5) | 5(10.0) | 0(0) | < 0.01 |
| Prior stroke, n(%) | 13(10.1) | 6(12.0) | 7(8.9) | 0.57 |
| COPD, n(%) | 13(10.1) | 4(8.0) | 9(11.4) | 0.53 |
| Pulmonary disease, n(%) | 17(13.2) | 5(10.0) | 12(15.2) | 0.39 |
| NYHA Functional Class III or IV, n(%) | 72(55.8) | 32(64.0) | 40(50.6) | 0.45 |
| STS score for MVP, % | 7.4(3.7-9.9) | 8.7(4.7-11.6) | 6.6(3.3-8.4) | 0.01 |
| STS score for MVR, % | 10.6(6.5-13.2) | 12.3(7.3-15.0) | 9.5(5.9-11.8) | 0.02 |

HT: hypertension, DL: dyslipidemia, DM: diabetes mellitus, AF/AFL: atrial fibrillation/atrial flutter, SHD: structure heart disease, COPD: chronic obstructive pulmonary disease, NYHA: New York Heart Association, STS score: The Society of Thoracic Surgeons Predicted Risk of Mortality score, MVP: mitral valve plasty, MVR: mitral valve replacement

**Table 2.**
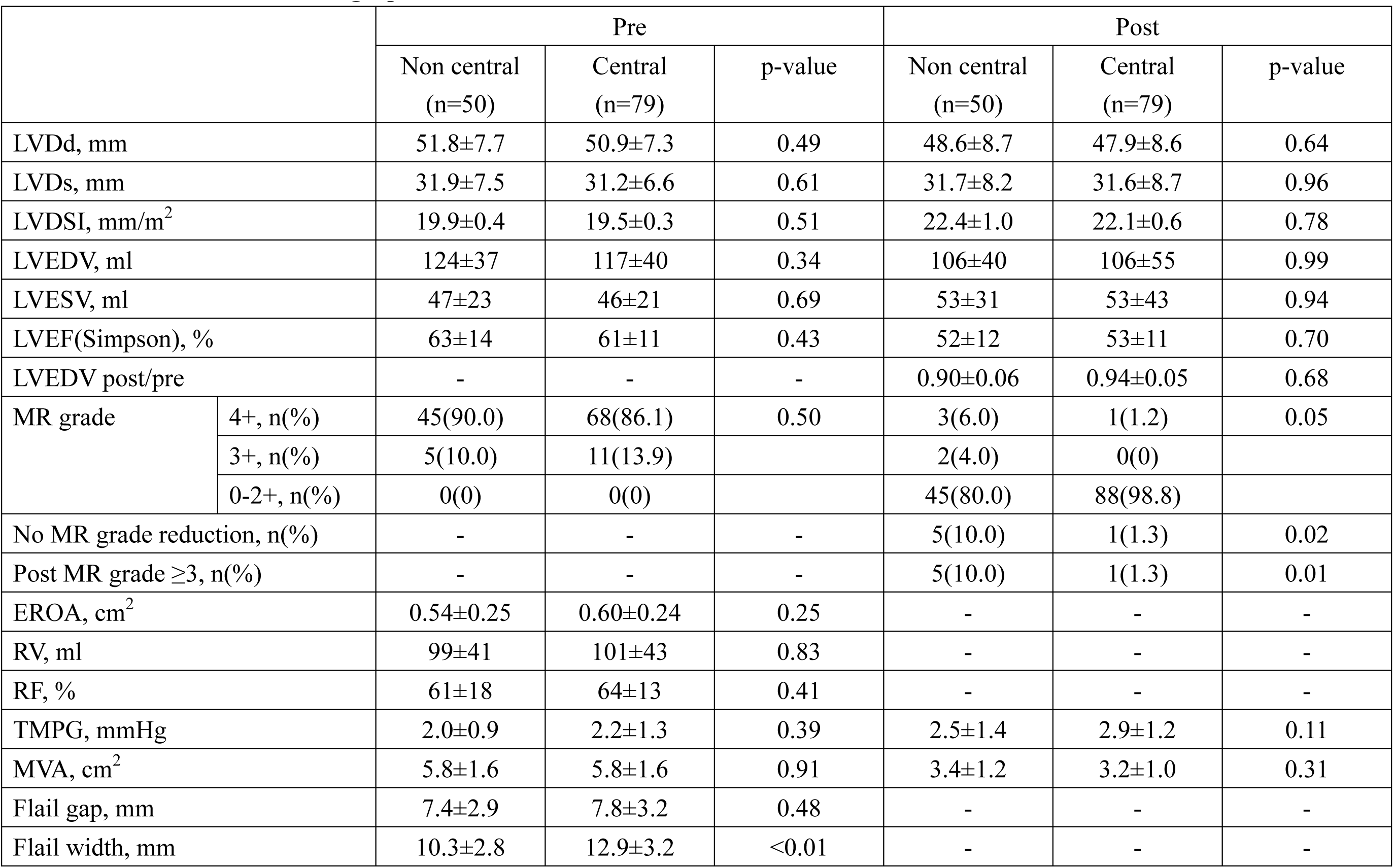
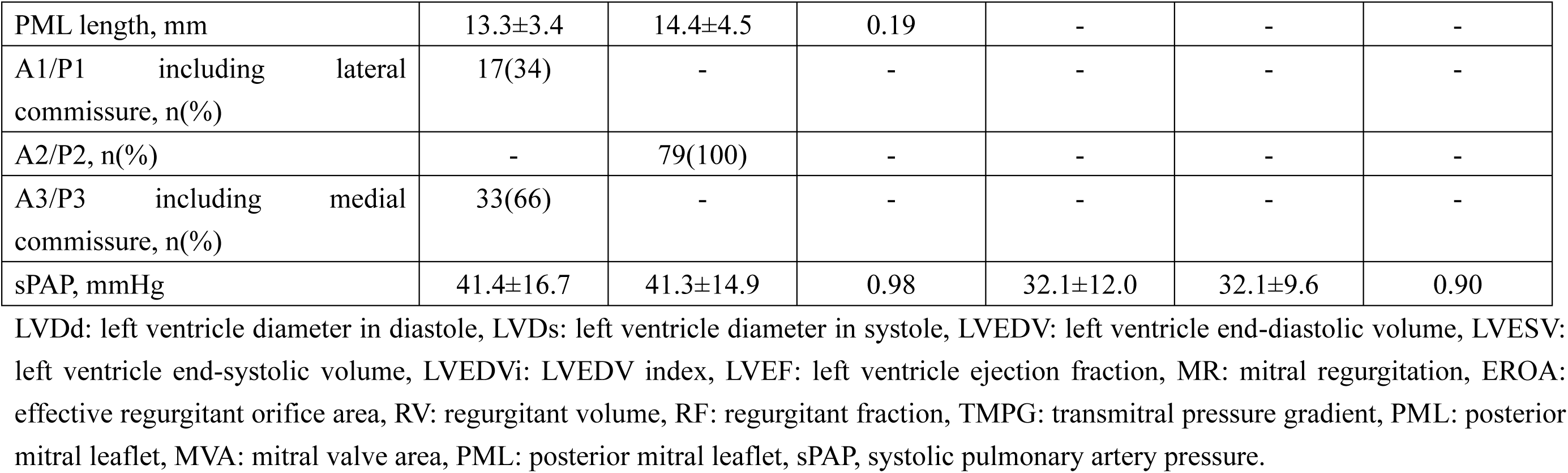
Procedural echocardiographic data.

|  |  | Pre |  |  | Post |  |  |
| --- | --- | --- | --- | --- | --- | --- | --- |
|  |  | Non central<br>(n=50) | Central<br>(n=79) | p-value | Non central<br>(n=50) | Central<br>(n=79) | p-value |
| LVDd, mm |  | 51.8±7.7 | 50.9±7.3 | 0.49 | 48.6±8.7 | 47.9±8.6 | 0.64 |
| LVDs, mm |  | 31.9±7.5 | 31.2±6.6 | 0.61 | 31.7±8.2 | 31.6±8.7 | 0.96 |
| LVDSI, mm/m<sup>2</sup> |  | 19.9±0.4 | 19.5±0.3 | 0.51 | 22.4±1.0 | 22.1±0.6 | 0.78 |
| LVEDV, ml |  | 124±37 | 117±40 | 0.34 | 106±40 | 106±55 | 0.99 |
| LVESV, ml |  | 47±23 | 46±21 | 0.69 | 53±31 | 53±43 | 0.94 |
| LVEF(Simpson), % |  | 63±14 | 61±11 | 0.43 | 52±12 | 53±11 | 0.70 |
| LVEDV post/pre |  | - | - | - | 0.90±0.06 | 0.94±0.05 | 0.68 |
| MR grade | 4+, n(%) | 45(90.0) | 68(86.1) | 0.50 | 3(6.0) | 1(1.2) | 0.05 |
|  | 3+, n(%) | 5(10.0) | 11(13.9) |  | 2(4.0) | 0(0) |  |
|  | 0-2+, n(%) | 0(0) | 0(0) |  | 45(80.0) | 88(98.8) |  |
| No MR grade reduction, n(%) |  | - | - | - | 5(10.0) | 1(1.3) | 0.02 |
| Post MR grade ≥3, n(%) |  | - | - | - | 5(10.0) | 1(1.3) | 0.01 |
| EROA, cm<sup>2</sup> |  | 0.54±0.25 | 0.60±0.24 | 0.25 | - | - | - |
| RV, ml |  | 99±41 | 101±43 | 0.83 | - | - | - |
| RF, % |  | 61±18 | 64±13 | 0.41 | - | - | - |
| TMPG, mmHg |  | 2.0±0.9 | 2.2±1.3 | 0.39 | 2.5±1.4 | 2.9±1.2 | 0.11 |
| MVA, cm<sup>2</sup> |  | 5.8±1.6 | 5.8±1.6 | 0.91 | 3.4±1.2 | 3.2±1.0 | 0.31 |
| Flail gap, mm |  | 7.4±2.9 | 7.8±3.2 | 0.48 | - | - | - |
| Flail width, mm |  | 10.3±2.8 | 12.9±3.2 | <0.01 | - | - | - |

|  |  |  |  |  |  |  |
| --- | --- | --- | --- | --- | --- | --- |
| PML length, mm | 13.3±3.4 | 14.4±4.5 | 0.19 | - | - | - |
| A1/P1 including lateral commissure, n(%) | 17(34) | - | - | - | - | - |
| A2/P2, n(%) | - | 79(100) | - | - | - | - |
| A3/P3 including medial commissure, n(%) | 33(66) | - | - | - | - | - |
| sPAP, mmHg | 41.4±16.7 | 41.3±14.9 | 0.98 | 32.1±12.0 | 32.1±9.6 | 0.90 |

LVDd: left ventricle diameter in diastole, LVDs: left ventricle diameter in systole, LVEDV: left ventricle end-diastolic volume, LVESV: left ventricle end-systolic volume, LVEDVi: LVEDV index, LVEF: left ventricle ejection fraction, MR: mitral regurgitation, EROA: effective regurgitant orifice area, RV: regurgitant volume, RF: regurgitant fraction, TMPG: transmitral pressure gradient, PML: posterior mitral leaflet, MVA: mitral valve area, PML: posterior mitral leaflet, sPAP, systolic pulmonary artery pressure.

The flail width was significantly narrower in NCpMR than in CpMR (10.3 ± 2.8 vs. 12.9 ± 3.2 mm, p < 0.01), while the posterior mitral leaflet (PML) length tended to be shorter in NCpMR (13.3 ± 3.4 vs. 14.4 ± 4.5 mm, p = 0.19). Other echocardiographic parameters were comparable between the 2 groups.

### Procedural characteristics and outcomes

Periprocedural characteristics and outcomes are presented in **Table 3**. There were no cases of in-hospital or 30-day mortality, procedural strokes, or major access-site complications in either group. Additionally, no instances of prolonged clip entanglement, clip embolization, or septal complications occurred. Among patients undergoing M-TEER, 3 cases (2 in NCpMR, one in CpMR) resulted in no MitraClip deployment due to the risk of SLDA. Post-procedurally, SLDA occurred in 4 NCpMR patients (8.0%), whereas no cases were observed in CpMR (p < 0.01). Residual MR grade 3+/4+ was identified in 6 patients (4.7%), with a significantly higher frequency in NCpMR than in CpMR (10.0% vs. 1.3%, p = 0.01). One NCpMR patient required emergent MV surgery within 24 hours due to SLDA but remained hemodynamically stable without circulatory support until surgery.

**Table 3.** Procedural outcomes and characteristics.

|  | All<br>(n=129) | Non central<br>(n=50) | Central<br>(n=79) | p-value |
| --- | --- | --- | --- | --- |
| In-Hospital Death, n(%) | 0(0) | 0(0) | 0(0) | 1.00 |
| 30-day mortality, n(%) | 0(0) | 0(0) | 0(0) | 1.00 |
| Procedural stroke, n(%) | 0(0) | 0(0) | 0(0) | 1.00 |
| Technical success, n(%) | 126(97.7) | 48(96.0) | 78(98.7) | 0.32 |
| Device success, n(%) | 118(91.4) | 40(80.0) | 78(98.7) | 0.03 |
| Procedural death, n(%) | 0(0) | 0(0) | 0(0) | 1.00 |
| Deployment failure, n(%) | 3(2.3) | 2(4.0) | 1(1.3) | 0.32 |
| MR at least moderate to severe, n(%) | 6(4.7) | 5(10.0) | 1(1.3) | 0.01 |
| SLDA following emergent surgery, n(%) | 1(0.7) | 1(2.0) | 0(0) | 0.17 |
| SLDA following elective surgery, n(%) | 3(2.3) | 3(6.0) | 0(0) | < 0.01 |
| Hospital length, day | 21.5±11.7 | 20.8±1.7 | 21.8±1.3 | 0.65 |
| Number of clips | 1.3±0.5 | 1.2±0.1 | 1.4±0.1 | 0.06 |
| Single clip, n(%) | 84(65.1) | 36(72.0) | 48(60.8) | 0.19 |
| MitraClip Generation 4, n(%) | 56(43.4) | 27(54.0) | 29(36.7) | 0.05 |
| Extended arm clips (XT/XTW), n(%) | 28(21.7) | 13(26.0) | 15(19.0) | 0.35 |
| Wide clips (NTW/XTW), n(%) | 27(20.9) | 6(12.0) | 21(26.6) | 0.04 |
| G2, n(%) | 73(56.6) | 23(31.5) | 50(68.9) | NA |
| G4NT, n(%) | 14(10.9) | 12(85.7) | 2(14.3) | NA |
| G4NTW, n(%) | 15(11.6) | 2(13.3) | 13(86.7) | NA |
| G4XT, n(%) | 18(14.0) | 12(66.7) | 6(33.3) | NA |
| G4XTW, n(%) | 9(7.0) | 1(11.1) | 8(88.9) | NA |

MR: mitral regurgitation, SLDA; single leaflet device attachment, G2: MitraClip generation second, G4: MitraClip generation forth.

Technical success was achieved in 122 patients (97.7%), with no significant difference between groups (96.0% vs. 98.7%, p = 0.32). However, device success was significantly lower in NCpMR than in CpMR (80.0% vs. 98.7%, p = 0.03). Although the number of implanted clips did not significantly differ between groups (1.2 ± 0.1 vs. 1.4 ± 0.1, p = 0.06), single-clip use was more frequent in NCpMR (72.0% vs. 60.8%, p = 0.19). The new-generation MitraClip G4 was used more often in NCpMR than in CpMR (54.0% vs. 36.7%, p = 0.05), while wide-type MitraClip (NTW/XTW) was significantly more common in CpMR (26.6% vs. 12.0%, p = 0.04). Residual MR grade 3+/4+ at discharge was significantly more common in NCpMR than in CpMR (10.0% vs. 1.3%, p = 0.01) (**Table 2** and **Figure 2**).

Compared to preoperative echocardiographic data, both groups exhibited evidence of LV reverse remodeling post-procedure, though the difference was not statistically significant (Left Ventricular End-Diastolic Volume (LVEDV) post-procedure / LVEDV pre-procedure; 0.90 ± 0.06 vs. 0.94 ± 0.05, p = 0.68).

### Worsening MR

One-year follow-up after M-TEER was completed in all patients. **Figure 3** presents MR grades at baseline, discharge, and follow-up assessments. Significant MR or reintervention was observed in 1.3% and 3.8% of CpMR cases at discharge and 1-month follow-up, respectively. In contrast, NCpMR had significantly higher rates, with 10.0% at discharge and 20.0% at 1 month. Between 6- and 12-month follow-up, significant MR in CpMR increased from 6.4% to 16.7%, whereas in NCpMR, it remained relatively stable at 23.9%. To further investigate these differences, **Figure 4** illustrates the proportion of patients with worsening MR (2+ and 3+/4+) after M-TEER, using the 1-month post-implantation MR grade as the baseline.

**Figure 3.**
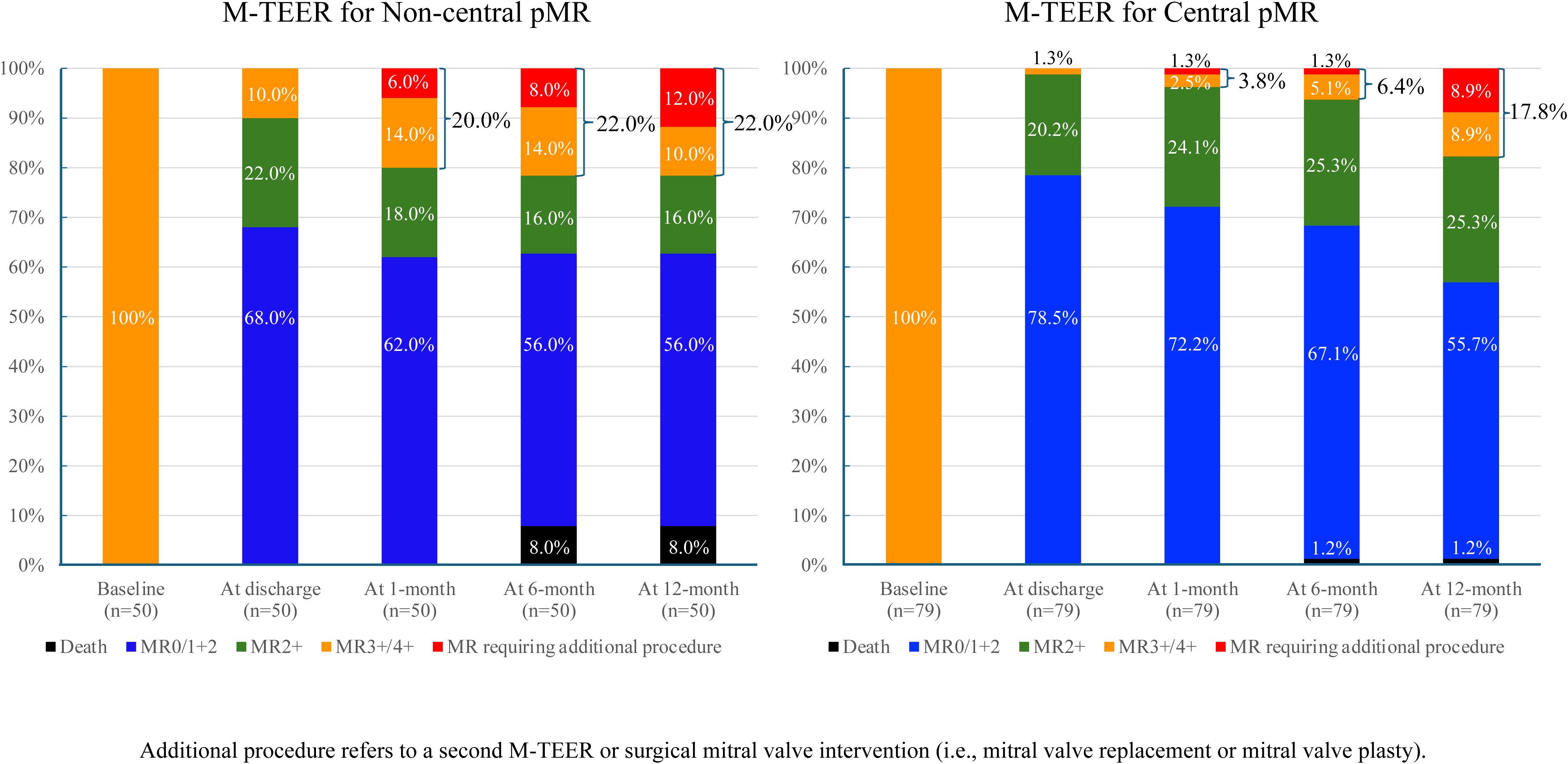
MR grade at baseline, discharge, and 1-, 6-, and 12-month following M-TEER among central and non-central groups. At discharge and 1 month, the rates of MR grade 3+/4+ and MR requiring intervention were 1.3% and 3.8% in the central group, compared to 10.0% and 20.0% in the non-central group, respectively. These rates were higher in the non-central group than in the central group. At 6 and 12 months, significant MR in the central group increased from 6.4% to 16.7%, while in the non-central group, it remained stable but higher at 23.9%. M-TEER = Transcatheter edge-to-edge mitral valve repair; pMR = primary mitral regurgitation.

**Figure 4.**
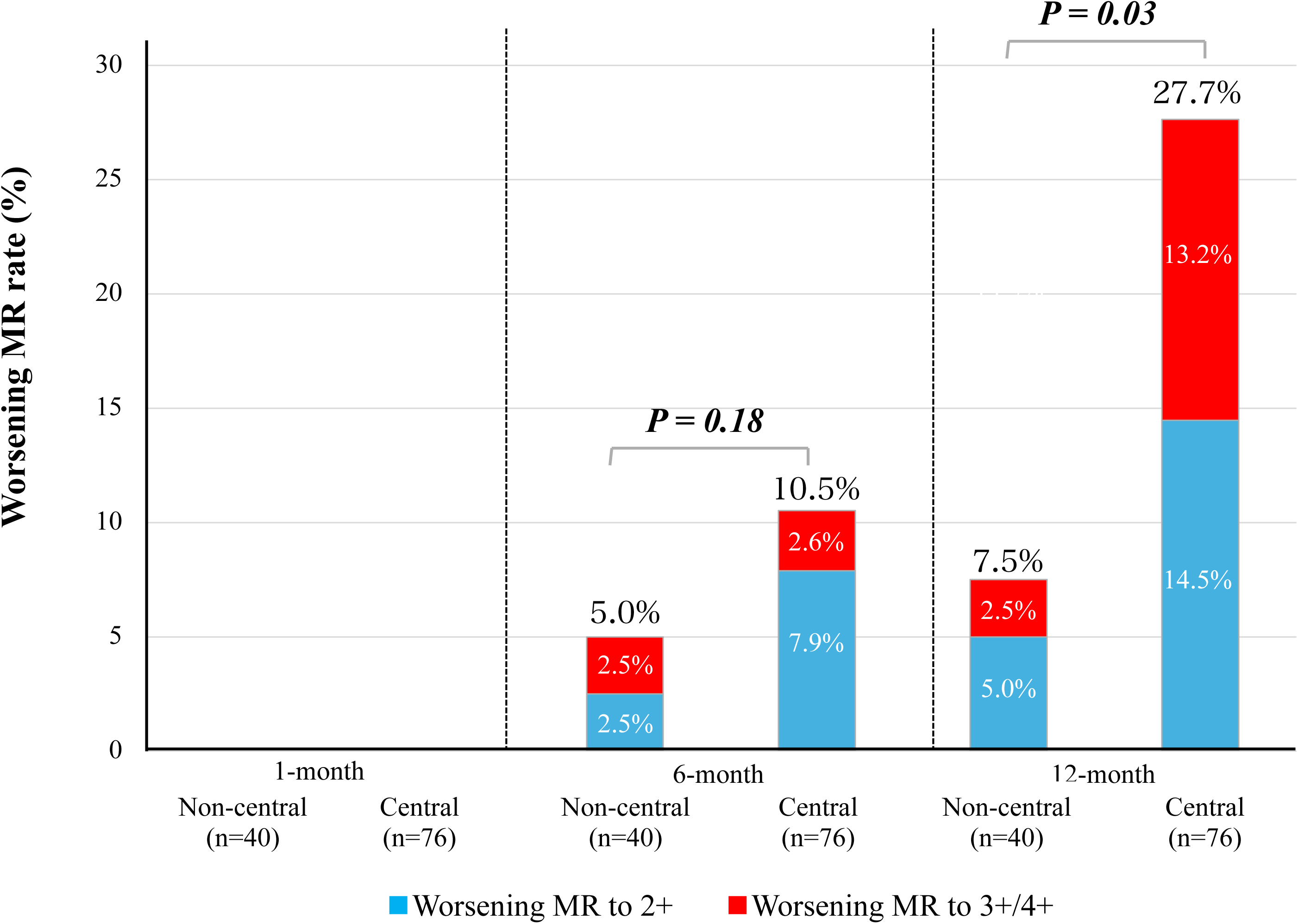
The proportion of patients with worsening to MR grade 2+ and MR 3+/4+ after M-TEER, using 1-month post-implantation MR as the baseline. At 6 months, MR worsening rates were 10.5% in the central group and 5.0% in the non-central group (p=0.18). At 12 months, worsening MR was significantly higher in the central group (27.7%) compared to the non-central group (7.5%, p=0.03). MR = mitral regurgitation.

At 6 months, worsening MR rates were 5.0% in NCpMR and 10.5% in CpMR (p = 0.18). At 12 months, rates were significantly lower in NCpMR compared to CpMR (7.5% vs. 27.7%, p = 0.03). When composite events were defined as deployment failure, SLDA, and residual MR grade 3+/4+, the 30-day freedom from composite events in NCpMR following M-TEER was 80%, which was lower than in CpMR (96%).

However, Kaplan-Meier analysis of composite events at 1 year showed no significant difference between groups (p = 0.16, **Figure 5**). To further investigate these findings, a 30-day landmark analysis was conducted to evaluate cumulative freedom from significant MR after M-TEER based on pMR lesion type. Surprisingly, and contrary to expectations, the analysis demonstrated that the incidence of residual MR grade 3+/4+ up to 1 year tended to be lower in NCpMR than in CpMR (p = 0.051, **Figure 6)**.

**Figure 5.**
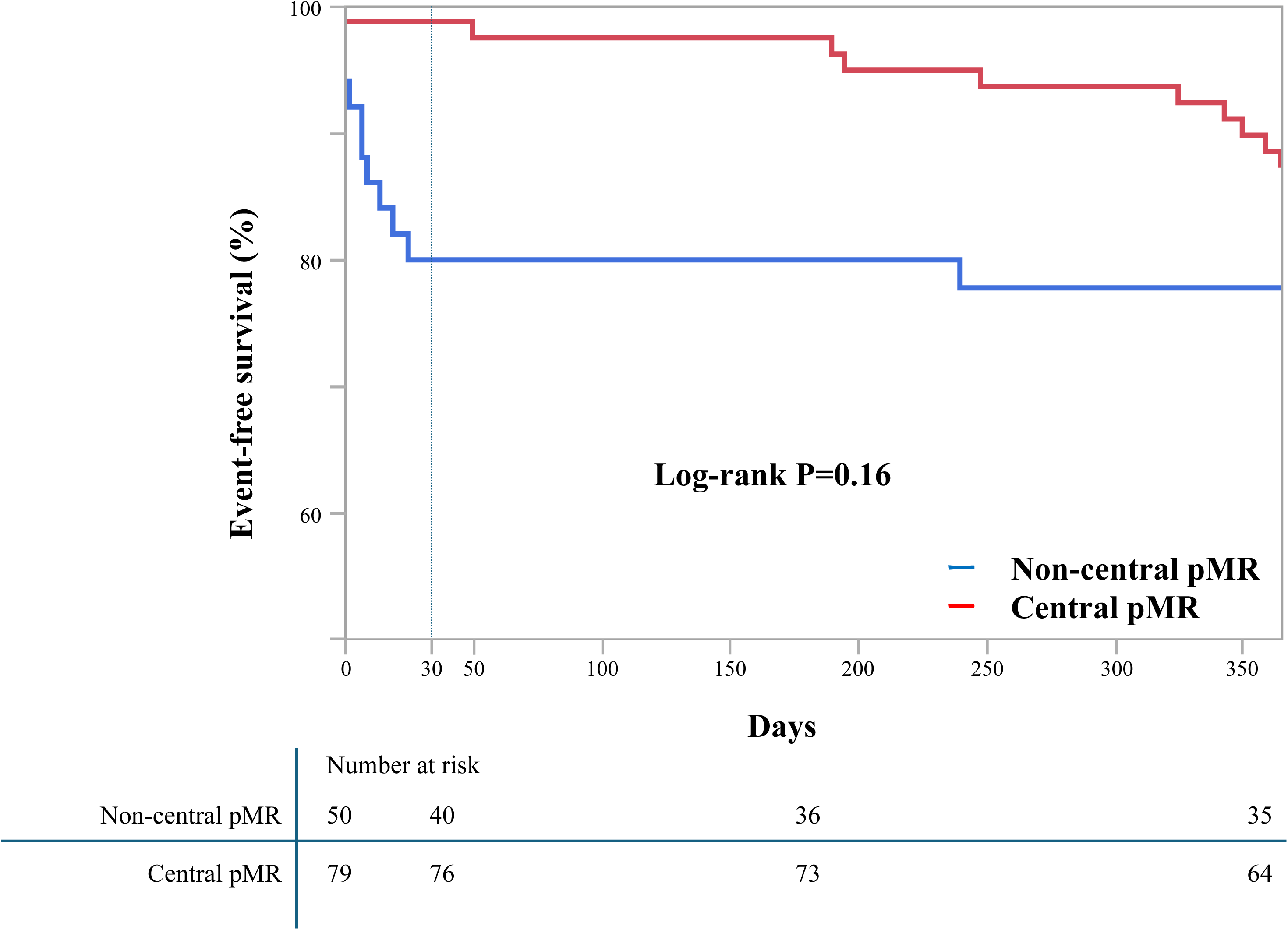
Cumulative free from composite events curves according to primary MR (pMR) lesion. Composite events were defined as deployment failure, SLDA, and residual MR grade 3+/4+. The 30-day cumulative freedom from composite events was significantly lower in non-central pMR (80%, blue curve) than in central pMR (96%, red curve) following M-TEER. However, Kaplan-Meier analysis at 1 year showed no significant difference (p=0.16) between groups. SLDA = single leaflet device attachment; M-TEER = transcatheter mitral edge-to-edge repair.

**Figure 6.**
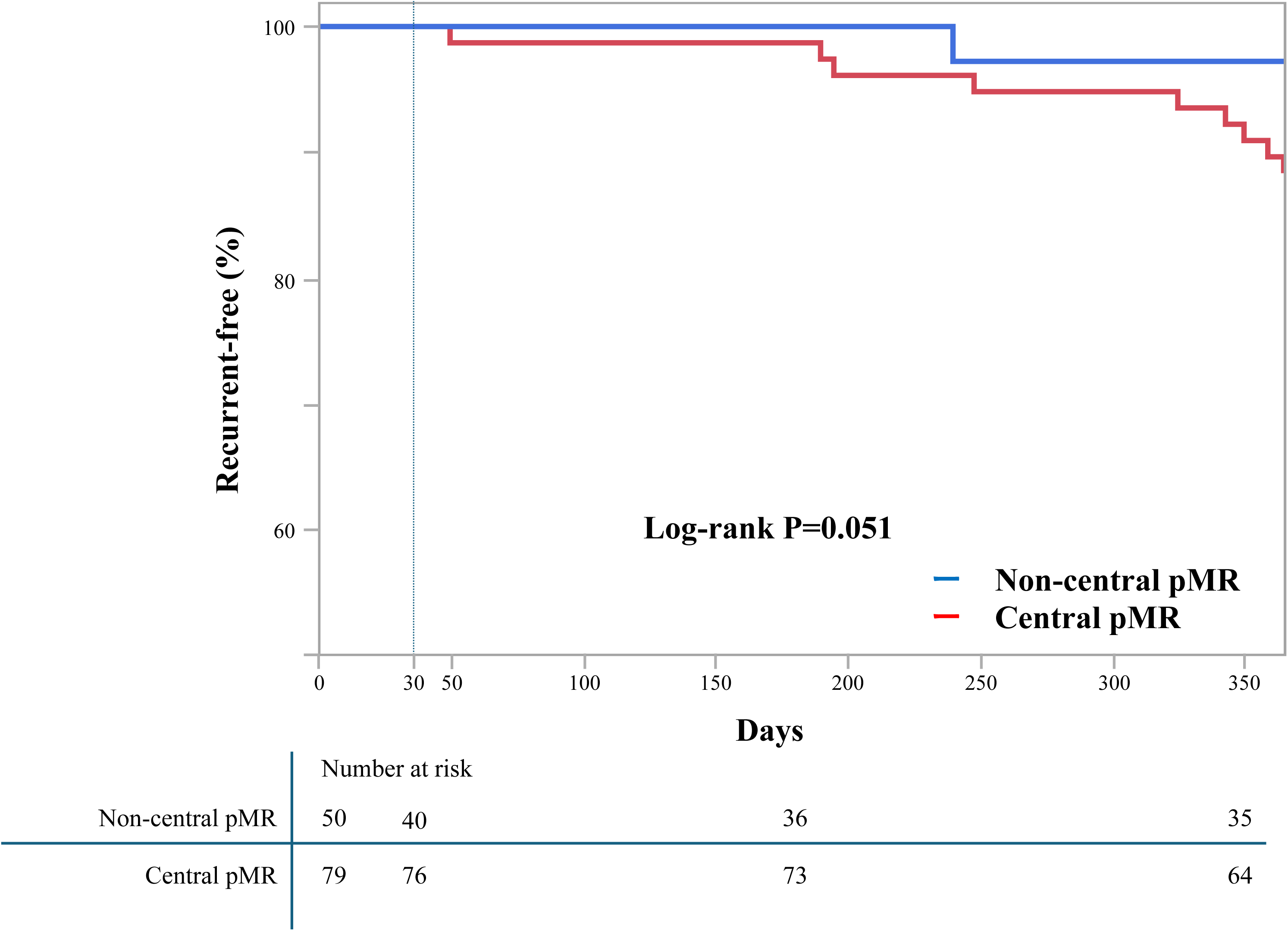
30-day landmark analysis of cumulative free from recurrent MR after M-TEER curves according to primary MR lesion. A 30-day landmark analysis of cumulative freedom from recurrent MR was performed after excluding patients with technical failure within 30 days. At 1 year, the incidence of recurrent MR grade 3+/4+ was higher in the central pMR group (red curve) compared to the non-central group (blue curve, p=0.051). pMR = primary mitral regurgitation; M-TEER = transcatheter edge-to-edge mitral valve repair; CI = confidence interval.

### Reintervention

The need for additional MV procedures due to recurrent MR was similar between groups [NCpMR: 1 (2.5%) vs. CpMR: 6 (7.7%), p = 0.23]. Among these 7 patients, one underwent surgical intervention, while the remaining 6 underwent a second M-TEER using MitraClip (**Table 4**). **Figure 7** presents a Sankey diagram illustrating MR severity changes over 1 year and outcomes for residual MR grade 3+/4+.

**Figure 7.**
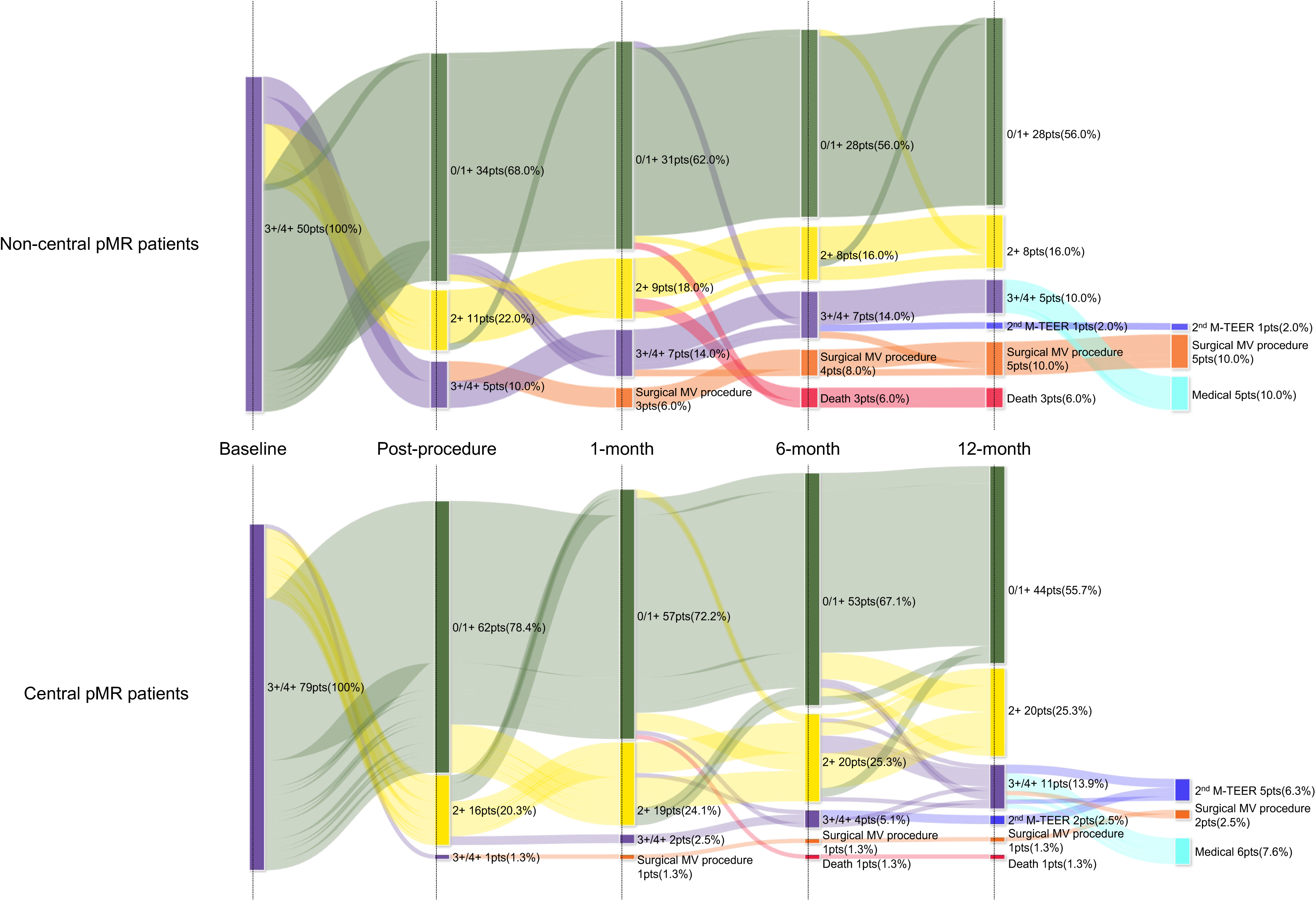
Sankey diagrams illustrating patient outcomes and MR grades at follow-ups after M-TEER. At 30 days, 10 NCpMR (10.0%) and 3 CpMR (3.8%) patients had residual MR grade 3+/4+. After one month, an additional 1 NCpMR (2.0%) and 10 CpMR (12.6%) experienced worsening MR. Among these patients: 5 NCpMR and 2 CpMR underwent surgical mitral valve procedures, 1 NCpMR and 5 CpMR underwent a second M-TEER, 5 NCpMR and 6 CpMR were managed conservatively

**Table 4.** Clinical outcomes and echocardiographic data at 12-month.

|  | All<br>(n=129) | Non central<br>(n=50) | Central<br>(n=79) | p-value |
| --- | --- | --- | --- | --- |
| 1-year mortality, n(%) | 5(4.2) | 4(10.0) | 1(1.2) | 0.04 |
| Recurrent MR 3+/4+ grade at 12-month after 1-month, n(%) | 12(10.2) | 1(2.5) | 11(13.9) | 0.03 |
| Additional MV procedure, n(%) | 7(5.4) | 1(2.5) | 6(7.7) | 0.23 |
| Surgical repair or replacement, n(%) | 1(0.8) | 0(0) | 1(1.3) | 0.36 |
| 2nd M-TEER, n(%) | 6(5.1) | 1(2.5) | 5(6.4) | 0.33 |

MR: mitral regurgitation, MV: mitral valve, M-TEER: transcatheter edge-to-edge mitral valve repair.

### One-year mortality and cause of death

At 1 year, all-cause mortality was observed in 4 patients from the NCpMR and one patient from the CpMR. However, only one death was attributed to cardiovascular causes (**Supplemental Table 2**). Notably, STS scores were generally higher in NCpMR compared to CpMR.

### Subgroup analysis of procedural complications based on MitraClip type

The incidence of complications by MitraClip type is presented in **Supplemental Table 3**. Three patients experienced clip deployment failure, and four patients had residual MR grade 3+/4+ at discharge. Notably, all of these cases involved the narrow and short clip arm type. In contrast, no instances of clip deployment failure or residual MR grade 3+/4+ at discharge were observed among patients treated with MitraClip NTW, XT, or XTW.

## Discussion

This study highlights three key findings. Non-central M-TEER demonstrated comparable safety to central M-TEER, acceptable efficacy in high surgical risk patients, and less worsening MR at 1 year compared to central M-TEER. To the best of our knowledge, this is the first report evaluating 1-year clinical and echocardiographic outcomes of M-TEER for non-central pMR in comparison to central pMR.

### Safety outcomes

There was no 30-day mortality in either group, and emergent intervention rates were similar, confirming M-TEER’s safety for central and non-central pMR. Although NCpMR had higher rates of residual MR grade 3+/4+, SLDA, and clip deployment failure, no 30-day mortality was observed. Even in the presence of residual MR or SLDA, emergent surgery rates remained comparable to CpMR, and all surgical patients remained hemodynamically stable until undergoing emergent or elective intervention. These findings reinforce M-TEER’s safety, aligning with the established safety profile of MitraClip, where suboptimal MR control has minimal impact on short-term clinical outcomes.

### Short-term efficacy

MR reduction of at least one grade was achieved in over 90% of patients in both groups, with 90% (45 cases) maintaining MR below grade 3+ in NCpMR. Although the incidence of residual MR grade 3+/4+ at discharge was higher in NCpMR than in CpMR, technical success rates of M-TEER were similar between groups, suggesting that M-TEER remains a highly feasible treatment for non-central MR, particularly in high surgical risk patients.

### Long-term efficacy

The rate of worsening MR at 1 year was significantly lower in NCpMR than in CpMR, as shown in **Figure 4** and **Figure 6**. These results suggest that when M-TEER successfully reduces MR initially, residual MR remains stable over time, particularly in NCpMR. Despite concerns regarding the management of non-central MR, our findings indicate effective MR control, further supporting M-TEER as a strong treatment option for high-surgical-risk patients.

Contrary to expectations, CpMR exhibited a higher rate of worsening MR and residual MR grade 3+/4+ at 1 year. The overall incidence of MR grade 3+/4+ at 1 year post-M-TEER in this study was 10.2%, aligning with previous studies^23^, confirming that our cohort does not represent an outlier in terms of MR control at 1 year. A prior study^13^ reported comparable MR control efficacy between NCpMR and CpMR up to 6 months post-procedure. However, in our study, MR worsening in CpMR became apparent between 6 months and 1 year, resulting in a significant difference between the groups.This is the first study to demonstrate the inferior long-term outcomes of CpMR compared to NCpMR, emphasizing the need for further investigation into the durability of M-TEER based on regurgitant jet location.

### Impact of device evolution on M-TEER outcomes

The G4 system introduced three new clip sizes: a longer clip arm type (XT), a wider clip arm type (NTW), and a longer and wider clip arm type (XTW). Wide clips (NTW/XTW) facilitate MR reduction with a single clip, particularly in cases with a large flail width. Additionally, the introduction of independent grasping and multiple clip sizes may have influenced the outcomes of this study. Independent grasping, in particular, may have contributed to easier leaflet capture in both central and non-central pMR. Regarding clip size, 56% of patients underwent M-TEER with the MitraClip G2. Device deployment failed in three patients, and four had residual MR grade 3+/4+ at discharge, all of whom received narrow, short-arm clips (**Supplemental Table 3**), specifically the MitraClip G2 and the narrow, short-arm type of MitraClip G4 NT. No such cases occurred with MitraClip G4 NTW/XT/XTW. Thus, short and narrow clips, including MitraClip G2 and MitraClip G4 NT, were suboptimal for treating both central and non-central pMR. Regarding the independent grasping mechanism, the introduction of independent grasping in the G4 system has enhanced the ability to securely grasp both leaflets, improving the feasibility of treating anatomically complex cases, particularly in large flail cases where simultaneous leaflet grasping is challenging. Increased use of MitraClip G4 may help reduce complications, including clip deployment failure and residual MR grade 3+/4+ at discharge.

### Proposal for the future based on this study: Initial M-TEER strategy for central and non-central pMR

Although CpMR exhibited a higher progression rate to significant MR, it remains treatable with a second M-TEER. In this study, 85.7% (6/7) of symptomatic recurrent MR cases in central pMR successfully underwent reintervention by M-TEER. In CpMR, particularly at the A2/P2 segment, MR often recurred from the same leaflet section where the clip was initially implanted. Our findings suggest that residual MR progression, especially between 6 months and 1 year, may result from unaddressed billowing or flail segments that continue to be subjected to LV pressure despite an apparent initial MR grade improvement. This scenario is especially relevant when the flail width is too broad to be fully addressed with a single clip. In such cases, although MR reduction may be achieved during the initial procedure, incomplete control of leaflet motion—such as persistent billowing—can become the source of recurrence (**Supplemental Video 1**). Therefore, even if MR appears to improve after one clip, if residual billowing remains, additional clips should be considered during the initial procedure to ensure durable leaflet coaptation and minimize the risk of recurrence. Non-central pMR in this study frequently recurred at the same leaflet section where the MitraClip was implanted, similar to central pMR—particularly from leaflet segments lateral to the clip in A1/P1 lesions and medial to the clip in A3/P3 lesions, often closer to the mitral leaflet margin (**Supplemental Video 2**). Unlike central pMR, performing a second M-TEER in non-central pMR is often unfeasible due to the limited available leaflet width for additional clip placement. As a result, most recurrent cases of non-central pMR in this study were managed either conservatively or with surgical intervention. Since M-TEER is primarily indicated for high surgical risk patients, those unable to undergo a percutaneous reintervention often have no viable surgical alternatives. Therefore, in the initial M-TEER for non-central pMR, it is crucial to place the clip as close to the leaflet edge as possible to prevent recurrence and ensure procedural durability. This strategy not only minimizes the risk of recurrence but also preserves space for potential future intervention if needed. Additionally, even in non-central pMR, cases with a wide flail gap should be considered for multiple clip placement from the outset to optimize leaflet coaptation and reduce the likelihood of recurrence.

### Study Limitation

The present analysis has some limitations. First, the small sample size may have limited the ability to detect differences in procedural and clinical outcomes. Additionally, the low event rate of procedural complications warrants cautious interpretation. Second, as a before-and-after study, the findings are subject to bias from temporal changes in clinical practice and operator learning curves. Third, patient selection may have been influenced by anatomical suitability for M-TEER, potentially introducing selection bias. Lastly, since September 2020, the MitraClip system transitioned from G2 to G4, with over half of cases using the older G2, which may have impacted technical and device success rates. Whether non-central M-TEER was an independent predictor of higher one-year mortality could not be statistically assessed due to the limited sample size. However, given the higher STS scores and prevalence of non-cardiac deaths in the non-central group, we do not consider M-TEER itself a contributing factor to increased mortality.

## Conclusions

Thirty-day mortality and perioperative emergent intervention rates for M-TEER were low and comparable between central and non-central pMR, confirming its safety. In non-central pMR, M-TEER achieved at least a one-grade MR reduction in 90% of patients, supporting its viability as a treatment option for inoperable cases. While MR grade at discharge was better in central pMR, MR worsening and progression to grade 3+/4+ were lower in non-central pMR. Notably, in anatomically challenging non-central pMR, once MR reduction was achieved, it was generally maintained long-term. These findings support M-TEER as a safe and effective treatment for high-risk patients with non-central pMR, highlighting its role in patients who are not candidates for surgical intervention.

## Data Availability

The data supporting the findings of this study are not publicly available due to patient privacy concerns, and therefore cannot be shared with readers.

## Acknowledgements

None.

## Abbreviations

MR: Mitral Regurgitation
MV: Mitral Valve
pMR: Primary Mitral Regurgitation
LV: Left Ventricular
M-TEER: Mitral Transcatheter Edge-to-Edge Repair
EVEREST: Endovascular Valve Edge-to-Edge Repair Study
US: United States
FDA: Food and Drug Administration
TTE: Transthoracic Echocardiography
TEE: Transesophageal Echocardiography
M-VARC: Mitral Valve Academic Research Consortium
SLDA: Single Leaflet Device Attachment
G2: second-generation (MitraClip system)
G4: fourth-generation (MitraClip system)
SD: standard deviation
IQR: interquartile range
CpMR: Central Primary Mitral Regurgitation
NCpMR: Non-Central Primary Mitral Regurgitation
NYHA: New York Heart Association
AF: Atrial Fibrillation
STS: Society of Thoracic Surgeons
RV: Regurgitant Volume
RF: Regurgitant Fraction
EROA: Effective Regurgitant Orifice Area
PML: Posterior Mitral Leaflet
LVEDV: Left Ventricular End-Diastolic Volume

---

**Supplemental Video 1.** A typical case of MR recurrence after M-TEER for central pMR.

**Supplemental Video 2**. A typical case of MR recurrence after M-TEER for non-central pMR.

